# ACCELEROMETER-BASED SEDENTARY BEHAVIOUR AND PHYSICAL ACTIVITY ARE ASSOCIATED WITH THE GUT MICROBIOTA IN 8507 INDIVIDUALS FROM THE POPULATION-BASED SCAPIS

**DOI:** 10.1101/2023.06.01.23290817

**Authors:** Gabriel Baldanzi, Sergi Sayols-Baixeras, Elin Ekblom-Bak, Örjan Ekblom, Koen F. Dekkers, Ulf Hammar, Diem Nguyen, Shafqat Ahmad, Ulrika Ericson, Daniel Arvidsson, Mats Börjesson, Peter J. Johanson, J Gustav Smith, Göran Bergström, Lars Lind, Gunnar Engström, Johan Ärnlöv, Beatrice Kennedy, Marju Orho-Melander, Tove Fall

## Abstract

**Objective:** Population-based studies investigating the relationship between physical activity and the gut microbiota composition have mainly relied on self-reported activity, potentially influenced by reporting bias. Here, we investigated associations of accelerometer-based sedentary behaviour and physical activity with the gut microbiota composition and functional profile in the large Swedish CArdioPulmonary bioImage Study.

**Methods:** In 8507 participants aged 50-65, the proportion of time in sedentary (SED), moderate-intensity (MPA), and vigorous-intensity (VPA) physical activity were estimated with hip-worn accelerometer. The gut microbiota was profiled using shotgun metagenomics of fecal samples. We fitted multivariable regression models, and adjusted for sociodemographic, lifestyle, and technical covariates while also accounting for multiple testing.

**Results:** Overall, SED and MPA were associated with microbiota species in opposite directions. For example, the strongest positive regression coefficient for MPA and the strongest negative for SED were with *Prevotella copri*, a plant-polysaccharide-degrading bacteria. Species associated with VPA aligned with the MPA associations, although with clear discrepancies. For instance, *Phocaeicola vulgatus* was negatively associated with MPA, while the association with VPA was non-significant and in the positive direction. Additional adjustment for dietary variables or adiposity attenuated some of the associations. For the functional profile, MPA and VPA were generally associated with lower capacity for amino acid degradation.

**Conclusion:** Our findings suggest that sedentary behaviour and physical activity are associated with a similar set of gut microbiota species and functions, but in opposite directions. Furthermore, the intensity of physical activity may have specific effects on certain species of the gut microbiota.

## INTRODUCTION

Physical activity has well-established health benefits, including reduced risk for cardiovascular disease, type 2 diabetes,^1,2^ and psychiatric conditions like depression.^3^ Conversely, sedentary behaviour, defined as sitting or non-sleep lying activities with low energy expenditure, is associated with increased risk of type 2 diabetes and cardiovascular mortality.^4–6^ Some studies have though indicated that the risks attributed to sedentary behaviour could be substantially attenuated by physical activity at high intensities.^7–9^

The gut microbiota is a community of microorganisms within the gastrointestinal tract that interacts with the host.^10^ Evidence suggests that the gut microbiota plays a role in the development of type 2 diabetes and cardiovascular diseases.^11–13^ Moreover, the gut microbiota may influence brain homeostasis through the microbiota-gut-brain axis, which includes microbe-produced neurotransmitters and other molecules that can affect the central nervous system via neuronal pathways or the immune system.^14^

Regular physical activity may affect the gut microbiota through various mechanisms, including modulation of the gut immune system, reduction in the intestinal transit time^15^ and splanchnic blood flow, transient increase in the intestinal permeability,^16^ and modulation of the enterohepatic circulation of bile acids.^17^ Smaller intervention studies in specific populations have reported changes to the gut microbiota composition after structured exercise, with a decrease in *Clostridium* and *Blautia* and an increase in *Bifidobacterium* and *Dorea*.^18–20^ In population-based studies, self-reported moderate and vigorous-intensity physical activity have been associated with higher gut microbiota diversity^21^, and sedentary behaviour with increased abundance of *Roseburia hominis* and *Erysipelatoclostridium* species.^22^ However, these studies had limited microbiota taxonomic resolution and used self-reported physical activity information, which may be affected by reporting bias.^23^ Therefore, there is a need for larger population-based studies that combine sensor-based physical activity assessment with gut microbiota profiled in higher taxonomic resolution. Here, we aimed to identify associations of accelerometer-based sedentary behaviour and physical activity with the gut microbiota analysed with deep shotgun metagenomics, in 8507 participants, using cross-sectional data from the Swedish CArdioPulmonary BioImage Study (SCAPIS).

## METHODS

### Study Population

The SCAPIS cohort includes 30 154 women and men aged 50 to 65 enrolled between 2013 and 2018 from six different regions in Sweden.^24^ Baseline investigation included wear of a hip-accelerometer for 7 days. Participants from Malmö and Uppsala were also invited to provide faecal samples for metagenomic analysis (n = 9831). The Swedish Ethical Review Authority approved the SCAPIS study (DNR 2010-228- 31M) and the present study (DNR 2018-315 with amendment 2020-06597). All participants provided written informed consent.

### Accelerometer data processing

Accelerometer data was processed using a previously described protocol^25^ with the added exclusion of registrations during estimated bedtime. For details, see supplemental methods. In brief, participants were instructed to wear the ActiGraph (Pensacola, USA) tri-axial accelerometer over the hip for 7 consecutive days, except during sleep and water-based activities. Raw data was transformed into counts per minute (cpm) over 60s epochs. Sedentary time (SED), low-intensity physical activity (LIPA), moderate-intensity physical activity (MPA), and vigorous-intensity physical activity (VPA) were defined as <200 cpm, 200 – 2689 cpm, 2690 – 6166 cpm, and ≥6167 cpm, respectively. Non-wear time was defined as periods of ≥60 min with zero counts, with intervals of maximum two minutes of 0 – 199 cpm. A valid day was defined as a day with >10 hours of wear time. We excluded 467 participants who had <4 valid days. The percentage of time in SED, MPA, or VPA were calculated by dividing the time spent in each activity by the total wear time.^25^ Because SED and LIPA were highly negatively correlated (Spearman’s correlation = −0.95, supplemental figure 1), we chose to exclude LIPA from the subsequent analyses.

### Faecal metagenomics

The gut microbiota was assessed through metagenomic analyses of faecal samples using a previously described protocol.^26^ In summary, faecal samples were collected at home, kept in the home freezer until the second study visit, and stored at −80°C until shipped to Clinical Microbiomics A/S (Copenhagen, Denmark) for DNA extraction, library preparation, sequencing with Ilumina Novaseq 6000 (Illumina, CA, USA), bioinformatics processing, and taxonomic annotation. Metagenomic species were defined by binning of co-abundance genes, as previously described.^27^ Alpha diversity, a metric of the diversity of species and the homogeneity of their abundance within a sample^28^ was estimated using the Shannon diversity index. Beta diversity, a metric of the composition dissimilarity between samples, was estimated using the Bray-Curtis dissimilarity.^29^ These estimations were performed with R package *vegan* on sequence data that was rarefied to 210 430 read-pairs for all samples to account for the differences in sequencing depth. The functional potential of the gut microbiota was defined by the abundance of genes of the manually curated gut metabolic modules^30^ (GMM) and microbiota-gut-brain modules^14^ (MGB). GMMs capture the microbiota metabolic potential and anaerobic fermentation capacity^30^, while MGBs comprise the capacity to degrade or produce potentially neuroactive compounds.^14^ To calculate the abundance of respective modules, we used the R package Omixer-RPM v.0.3.2^31^ considering a minimum module coverage of 66.6%. Analyses were focused on species identity, GMMs, and MGBs with a relative abundance >0.01% in ≥1% of individuals, resulting in 1336 species, 76 GMMs, and 28 MGBs for further analysis. Before statistical analysis, species, GMM, and MGB were log(x+1) transformed, where x denotes the relative abundance. For details, see supplemental methods.

### Covariates

Covariate information was obtained from the SCAPIS questionnaire, and anthropometric measurements and fasting plasma samples collected during study site visit. Mean daily intake of alcohol, fibre, added sugar, protein, carbohydrate, fat, and total energy were estimated from the food frequency questionnaire.^32,33^ Fibre, protein, carbohydrate, fat, and added sugar intake were transformed to percentages of non-alcohol energy intake. We categorized smoking status as current, former, or non-smoker, and highest achieved education level as incomplete compulsory, complete compulsory, secondary, or university education. Country of birth was grouped as Scandinavia (Sweden, Denmark, Norway, or Finland), non-Scandinavian Europe, Asia, and other countries. Information on medications prescribed for hypertension, type 2 diabetes, dyslipidaemia, depression, and anxiety within six months before the first site visit was retrieved from the Swedish Prescribed Drug Register (supplemental methods). Proton-pump inhibitor usage was defined as a measurable level of omeprazole or pantoprazole metabolites in plasma. Antibiotic use was defined as a dispensed prescription (Anatomical Therapeutic Chemical code J01) up to three months before the first visit.

### Statistical method

#### Model specification

Based on current literature^21,25,34^, we created a directed acyclic graph prior to the analysis phase using the DAGitty tool (www.daggity.net, supplemental figure 2), and selected potential confounders for the main model based on the d-separation criteria.^35^ The main model comprised age, sex, alcohol intake, smoking, education, country of birth, study site, and month of accelerometer wear. We also adjusted for technical variation covariates including total wear time, percentage of wear time on weekend, and faecal DNA extraction plate. To identify associations independent of dietary intake or adiposity, we explored two additional models: (1) Diet-adjusted model: additional adjustment for total energy intake and intake from added sugars, proteins, carbohydrates, and fibre, or (2) BMI/WHR-adjusted model: additional adjustment for body mass index (BMI) and waist-hip ratio (WHR). Statistical analyses were conducted in R version 4.1.1 (http://www.r-project.org).

#### Alpha and beta diversity

For alpha diversity, we applied linear regression models with Shannon index as the dependent variable and SED, MPA, and VPA separately as independent variables, while adjusting for covariates. Regression coefficients presented represent standard-deviation changes in SED, MPA, or VPA. For beta diversity, we used distance-based redundancy analysis to estimate the proportion of the interindividual gut microbiota dissimilarity explained by SED, MPA, and VPA. The analyses accounted for the covariates by using the parameter “condition” in the function “capscale” (R package *vegan*) and p-values were calculated based on 9999 permutations.

#### Species, gut metabolic modules, and microbiota-gut-brain modules

To identify microbiota features associated with physical activity overall, we applied a series of linear regression models with each species, GMM, and MGB introduced as the dependent variable, and SED, MPA, and VPA jointly introduced as independent variables together with the main model covariates. We used the likelihood ratio test to compare these models to an alternative model that only contained the covariates. To control for multiple testing, we applied the Benjamini-Hochberg method with a 5% false discovery rate and reported significance as q-values.^36^ The species, GMMs, and MGBs identified (q-value <0.05) where subsequently evaluated in models where SED, MPA, and VPA were introduced separately as an independent variable while adjusting for main model covariates, as well as for the diet-adjusted model and the BMI/WHR-adjusted model. To identify associations potentially caused by single influential observations, we calculated dfbetas for the main model. If removing the observation with the highest dfbeta resulted in a change in the direction of the association or a p-value ≥0.05, we discarded the association. Lastly, remaining species were examined in two sensitivity analyses: (1) additional adjustment for use of proton-pump inhibitors and medication for hypertension, type 2 diabetes, dyslipidaemia, anxiety, and/or depression; (2) exclusion of participants that were prescribed antibiotic treatment within the last three months.

## RESULTS

### Study population

The study population consisted of 4197 participants from Malmö and 4310 participants from Uppsala with valid accelerometer data, faecal metagenomics data, and complete information on the main model covariates (Table 1).

**Table 1.** Characteristics of the study population stratified by study site.

|  | All | Malmö | Uppsala |
| --- | --- | --- | --- |
|  | N=8507 | N=4197 | N=4310 |
| **Age, years** | 57.6 [53.8;61.4] | 57.4 [53.7;61.2] | 57.8 [53.9;61.5] |
| **Women, N (%)** | 4533 (53.3%) | 2278 (54.3%) | 2255 (52.3%) |
| **Accelerometer:** |  |  |  |
| **Wear time in sedentary, %** | 55.5 [48.5;62.0] | 55.6 [48.7;62.3] | 55.4 [48.4;61.7] |
| **Wear time in moderate-intensity, %** | 4.7 [3.2;6.6] | 4.3 [2.8;6.3] | 5.1 [3.6;6.9] |
| **Wear time in vigorous-intensity, %** | 0.06 [0.00;0.48] | 0.03 [0.00;0.37] | 0.10 [0.02;0.61] |
| **Valid days, days** | 7 [6;7] | 7 [6;7] | 7 [7;7] |
| **Wear time/day, h** | 14.5 [13.7;15.4] | 14.1 [13.3;14.8] | 15.0 [14.1;16.0] |
| **Wear time registered during weekends, %** | 28.6 [28.6;28.6] | 28.6 [28.6;28.6] | 28.6 [28.6;28.6] |
| **Alcohol intake, g/day** | 5.4 [1.8;10.3] | 5.2 [1.6;10.3] | 5.6 [1.9;10.2] |
| **Smoking, N (%):** |  |  |  |
| **Never** | 4379 (51.5%) | 1834 (43.7%) | 2545 (59.0%) |
| **Former** | 3046 (35.8%) | 1650 (39.3%) | 1396 (32.4%) |
| **Current** | 1082 (12.7%) | 713 (17.0%) | 369 (8.6%) |
| **Education, N (%):** |  |  |  |
| **Compulsory<sup>a</sup>** | 784 (9.2%) | 462 (11.0%) | 322 (7.5%) |
| **Upper secondary** | 3790 (44.6%) | 1998 (47.6%) | 1792 (41.6%) |
| **University** | 3933 (46.2%) | 1737 (41.4%) | 2196 (51.0%) |
| **Country of birth, N (%):** |  |  |  |
| **Scandinavia<sup>b</sup>** | 7183 (84.4%) | 3308 (78.8%) | 3875 (89.9%) |
| **Europe** | 752 (8.8%) | 584 (13.9%) | 168 (3.9%) |
| **Asia** | 382 (4.5%) | 210 (5.0%) | 172 (4.0%) |
| **Other** | 190 (2.2%) | 95 (2.3%) | 95 (2.2%) |
| **BMI and WHR** | N=8503 | N=4193 | N=4310 |
| **BMI, kg/m<sup>2</sup>** | 26.4 [24.0;29.4] | 26.6 [24.1;29.6] | 26.3 [24.0;29.1] |
| **Waist-hip ratio** | 0.9 [0.9;1.0] | 0.9 [0.8;1.0] | 0.9 [0.9;1.0] |
| **Dietary variables** | N=8416 | N=4144 | N=4272 |
| **Total energy intake (kcal/day)** | 1594 [1239;2049] | 1572 [1221;2059] | 1611 [1268;2040] |
| **Protein, E%** | 16.6 [14.7;18.6] | 16.5 [14.5;18.7] | 16.7 [15.0;18.6] |
| **Carbohydrate, E%** | 43.8 [39.4;48.0] | 43.9 [39.2;48.2] | 43.8 [39.5;47.9] |
| **Fat, E%** | 37.2 [33.3;41.1] | 37.3 [33.3;41.3] | 37.1 [33.3;40.8] |
| **Fibre, E%** | 2.3 [1.8;2.9] | 2.3 [1.8;2.9] | 2.4 [1.9;2.9] |
| **Added Sugar, E%** | 7.3 [4.9;10.6] | 7.3 [4.8;10.9] | 7.2 [5.1;10.3] |
| **Sensitivity analysis – medication use** | N=7560 | N=3295 | N=4265 |
| **Type 2 diabetes, N (%)** | 301 (4.0%) | 145 (4.4%) | 156 (3.7%) |
| **Hypertension, N (%)** | 1823 (24.1%) | 830 (25.2%) | 993 (23.3%) |
| **Dyslipidaemia, N (%)** | 798 (10.6%) | 380 (11.5%) | 418 (9.8%) |
| **Depression, N (%)** | 786 (10.4%) | 342 (10.4%) | 444 (10.4%) |
| **Anxiety, N (%)** | 278 (3.7%) | 143 (4.3%) | 135 (3.2%) |
| **Proton pump inhibitors, N (%)** | 285 (3.8%) | 155 (4.7%) | 130 (3.0%) |
| **Sensitivity analysis – antibiotic use** | N=8507 | N=4197 | N=4310 |
| **Antibiotic last 3 months, N (%)** | 498 (5.9%) | 270 (6.5%) | 228 (5.3%) |

Continuous variables described as median and 25<sup>th</sup> and 75<sup>th</sup> percentiles. Categorical variables described as absolute numbers and percentages. WHR: Waist-hip ratio. <sup>a</sup> Incomplete or complete compulsory education. <sup>b</sup> Sweden, Denmark, Finland or Norway. E%: percentages of non-alcohol energy intake.

### Sedentary behaviour and physical activity associations with alpha diversity and beta diversity

We found that SED was associated with lower alpha diversity (β = −0.020, p-value = 1.2×10^-5^), whereas MPA and VPA were associated with higher alpha diversity (MPA: β =0.024, p-value = 5.5×10^-8^; VPA: β = 0.032, p-value = 1.7×10^-12^). Similar results were found in the diet-adjusted model. In the BMI/WHR-adjusted model, only MPA and VPA were associated with alpha diversity (MPA: β = 0.017, p-value = 1.3×10^-4^; VPA: β = 0.022, p-value = 1.4×10^-6^).

To evaluate the contribution of sedentary behaviour and physical activity on the interindividual variation of the gut microbiota composition, we performed distance-based redundancy analysis on the Bray-Curtis dissimilarly matrix. After conditioning on the main model covariates, the adjusted R^2^ for SED was 0.083%, for MPA 0.084%, and for VPA 0.081%. The adjusted R^2^ for SED, MPA and VPA jointly was 0.188% in the main model, 0.141% in the diet-adjusted model, and 0.154% in the BMI/WHR-adjusted model (all p-values = 1×10^-4^). For comparison, the R^2^ for BMI without accounting for other variables was 0.76%, for fibre intake 0.39%, and for the dietary variables combined 0.69%.

### Sedentary behaviour and physical activity were associated with a large number of the species present in the human gut

We identified 345 species associated with physical activity (supplemental table 1). Among these, SED was associated with 224 species (q-value <0.05), MPA with 245 species, and VPA with 228 species. However, we found that six associations for SED, 16 associations for MPA, and 31 associations for VPA were caused by single influential observations (supplemental table 2). Overall, associations for SED and MPA mirrored each other (figure 1). The Pearson correlation between the main model regression coefficients for SED and MPA was −0.92, while between SED and VPA was −0.67. Amongst the largest coefficients, SED was positively, and MPA and VPA were negatively associated with *Blautia obeum* (unique identifier HG3A.0001) and *Ruminococcus torques*. Additionally, SED was negatively and MPA was positively associated with *Prevotella copri* and an unclassified *Eubacteriales* species (identifier HG3A.0100). The largest positive coefficients for VPA were with four unclassified *Eubacteriales* species (HG3A.0125, HG3A.0100, HG3A.0062, HG3A.0162).

**Figure 1.**
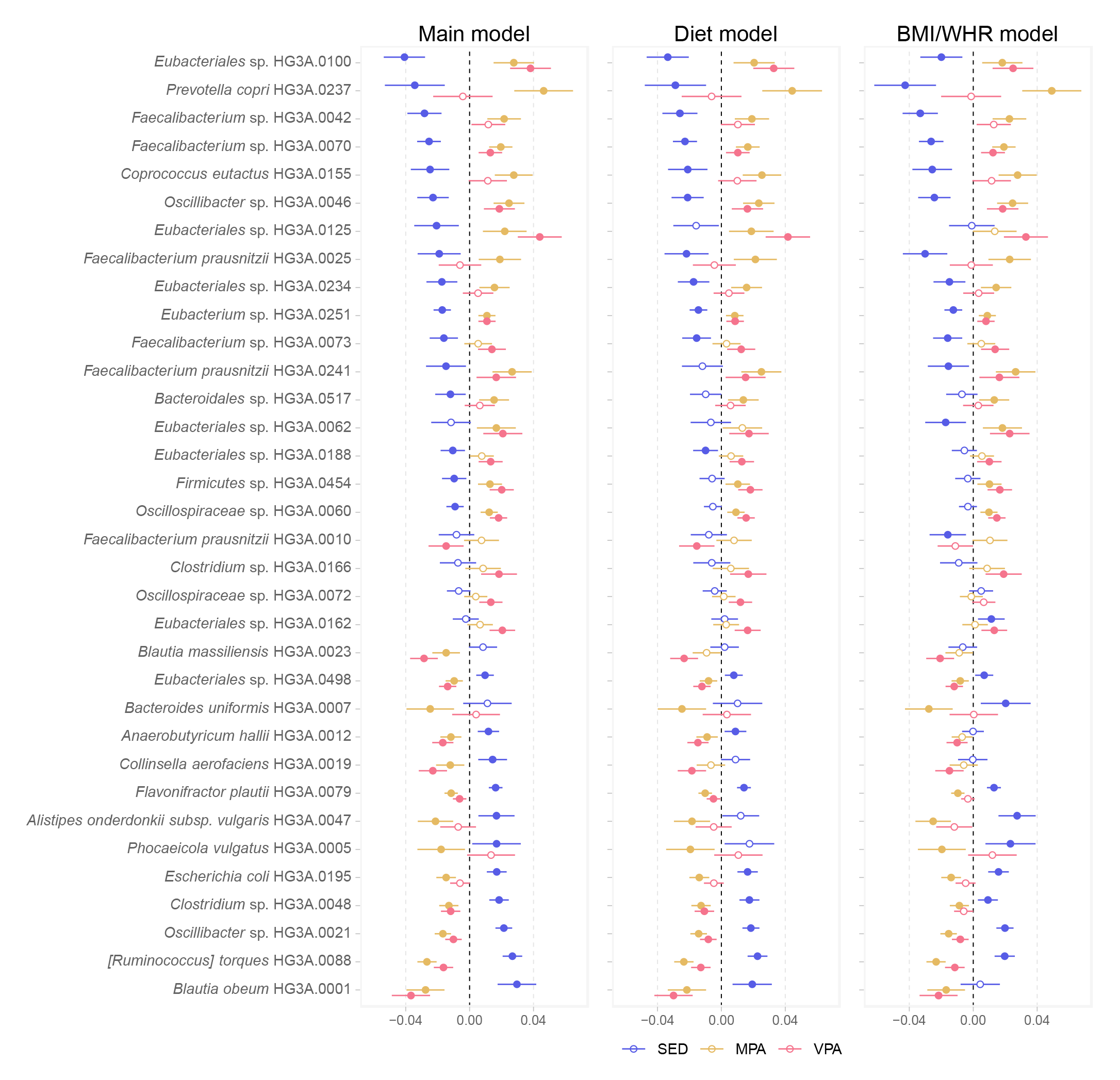
Top 20 associations based on the absolute regression coefficients for SED, MPA, and VPA with gut microbiota species. Effect estimates show changes in the log(relative abundance+1) of species by standard-deviation changes in SED, MPA, or VPA. Main model: adjustment for age, sex, alcohol intake, smoking, education, country of birth, study site, month of accelerometer wear, total accelerometer wear time, percentage of wear time on weekend, and fecal DNA extraction plate. Diet model: main model with additional adjustment for total energy intake, and percentage of energy intake from carbohydrates, protein, fibers, and added sugars. BMI/WHR model: main model with additional adjustment for BMI and waist-hip ratio. Circles are the regression coefficients and bars represent the 95% confidence intervals. Filled circles are associations with q-values <0.05. SED: percentage of time in sedentary behaviour; MPA: percentage of time in moderate-intensity physical activity; VPA: percentage of time in vigorous-intensity physical activity.

Regression coefficients for MPA and VPA were in largely consistent in direction and magnitude (Pearson correlation = 0.67). However, for specific species, there were marked differences in the association with MPA or VPA. For instance, *P. copri* and *Faecalibacterium prausnitzii* (HG3A.0025) were among the largest positive coefficients for MPA, but were not associated with VPA. Other differences include *Phocaeicola vulgatus* and *Bacteroides uniformis*, which were negatively associated with MPA, but not with VPA (figure 1).

The main model and the diet-adjusted model regression coefficients were highly correlated (Pearson correlation >0.99 for SED, MPA, and VPA), although generally attenuated in diet-adjusted model (supplemental table 2 and supplemental figure 3). The BMI/WHR-adjusted model also produced similar coefficients for MPA and VPA compared to the main model coefficients (Pearson correlation >0.97). However, regression coefficients for SED from the BMI/WHR model and from the main model were not as strongly correlated (Pearson correlation = 0.89). Certain main model associations for SED were substantially attenuated in the BMI/WHI model, especially for *B. obeum* (HG3A.0001) which was no longer associated with SED (figure 1 and supplemental table 2). On the other hand, the negative association between SED and *F. prausnitzii* (HG3A.0025) became stronger in the BMI/WHR model.

To investigate the potential effect of medication usage on the associations identified in the main model (q-value <0.05), we performed further adjustment for use of proton-pump inhibitors and medications for hypertension, type 2 diabetes, dyslipidaemia, anxiety, and/or depression. After this adjustment, SED was associated (p-value < 0.05) with 187 of the 224 species associated in the main model, MPA with 205 of 245, and VPA with 169 of 228 species. In the sensitivity analysis removing 498 participants who had used any antibiotic in the last 3 months, SED was associated (p-value < 0.05) with 215 species, MPA with 221, and VPA with 195 (supplemental table 3). The direction of the associations did not change.

### Sedentary behaviour and physical activity are associated with the gut microbiota functional potential

The functional potential was determined by the abundance of previously curated functional modules, which include modules of carbohydrate, amino acid, or lipid degradation,^30^ and modules of the microbiota-gut-brain axis.^14^ We identified 74 functional modules associated with physical activity (supplemental table 4). Out of 17 modules of carbohydrate degradation, SED was associated with lower or higher abundance of 12 modules in the main model (figure 2 and supplemental table 5). The strongest negative association was between SED and degradation of arabinoxylan, which is a dietary fibre. Even in the diet-adjusted model, SED was associated with degradation of arabinoxylan. Out of the 26 modules of amino acid degradation, SED was associated with higher abundance of 17 in the main model. In the diet-adjusted model, SED was associated with higher abundance of 15 modules of amino acid degradation, whereas MPA was associated with lower abundance of 14, and VPA with lower abundance of 7.

**Figure 2.**
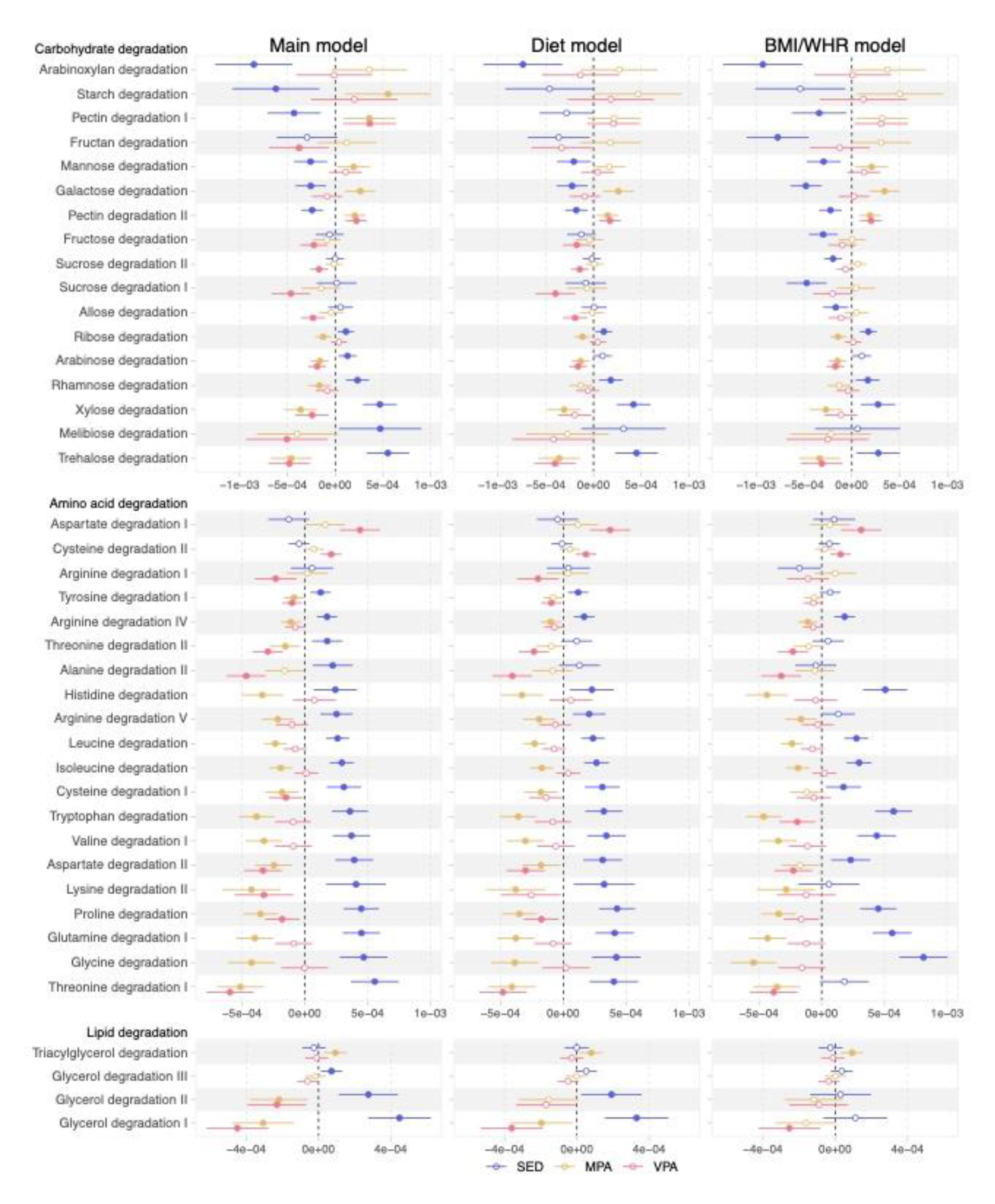
Associations of SED, MPA and VPA with functional modules of carbohydrate, amino acid, and lipid degradation. Effect estimates show changes in the log(relative abundance+1) of modules by standard-deviation change in SED, MPA, or VPA. Main model: adjustment for age, sex, alcohol intake, smoking, education, country of birth, study site, month of accelerometer wear, total accelerometer wear time, percentage of wear time on weekend, and fecal DNA extraction plate. Diet model: main model with additional adjustment for total energy intake, and percentage of energy intake from carbohydrates, protein, fibers, and added sugars. BMI/WHR model: main model with additional adjustment for BMI and waist-hip ratio. Circles are the regression coefficients and bars represent the 95% confidence intervals. Filled circles are associations with q-values <0.05. SED: percentage of time in sedentary behaviour; MPA: percentage of time in moderate-intensity physical activity; VPA: percentage of time in vigorous-intensity physical activity.

Among the modules of the microbiota-gut-brain axis, the largest main model coefficient was found for heat-shock protein ClpB, which was negatively associated with SED, and positively associated with MPA and VPA (figure 3 and supplemental table 5). Comparing main model and BMI/WHR-adjusted model associations, we observed marked differences for the butyrate synthesis I module. In the main model, VPA was positively associated with butyrate synthesis I, while no association was detected for SED or MPA. In the BMI/WHR-adjusted model, we could not detect the association for VPA; instead, SED was positively and MPA was negatively associated with this module. No association was driven by a single influential observation.

**Figure 3.**
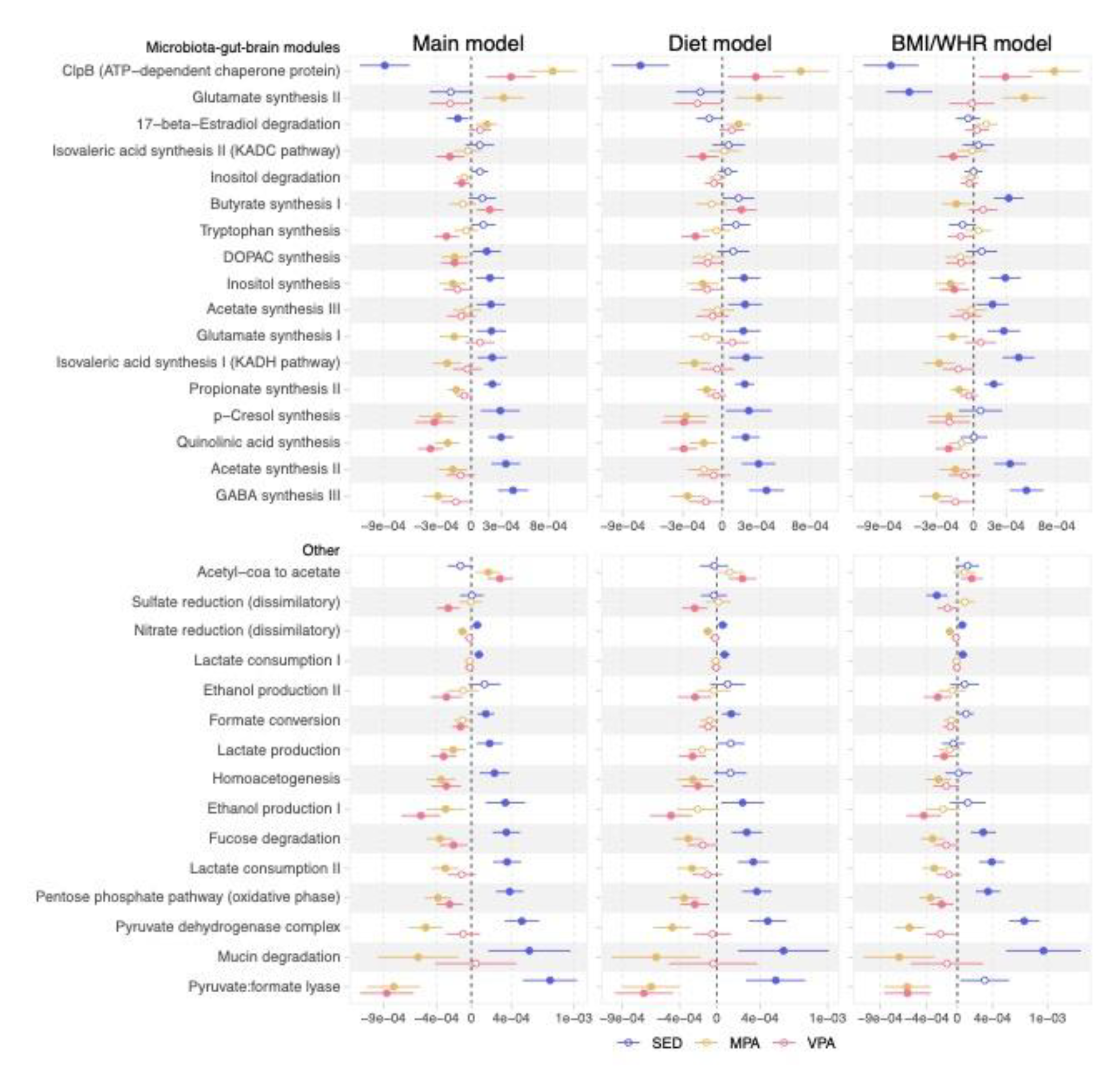
Associations of SED, MPA and VPA with microbiota-gut-brain modules and other functional modules. Effect estimates show changes in the log(relative abundance+1) of modules by standard-deviation change in SED, MPA, or VPA. Main model: adjustment for age, sex, alcohol intake, smoking, education, country of birth, study site, month of accelerometer wear, total accelerometer wear time, percentage of wear time on weekend, and fecal DNA extraction plate. Diet model: main model with additional adjustment for total energy intake, and percentage of energy intake from carbohydrates, protein, fibers, and added sugars. BMI/WHR model: main model with additional adjustment for BMI and waist-hip ratio. Circles are the regression coefficients and bars represent the 95% confidence intervals. Filled circles are associations with q-values <0.05. SED: percentage of time in sedentary behaviour; MPA: percentage of time in moderate-intensity physical activity; VPA: percentage of time in vigorous-intensity physical activity.

## DISCUSSION

In this largest-to-date population-based study of physical activity with gut microbiome encompassing 8507 individuals, we found that, from the 1336 species investigated, 345 (25.8%) were associated with physical activity. Overall, SED and MPA were associated with the same set of species but with regression coefficients in opposite directions. Furthermore, MPA and VPA had concordant associations with gut microbiota species, although with notable exceptions. Similar results were observed after adjustment for dietary variables, but some associations for SED were markedly affected by the adjustment for BMI/WHR. Additionally, SED, MPA, and VPA were associated with specific microbial functions, some potentially involved in the microbiota-gut-brain axis.

### The associations of sedentary behaviour and physical activity with gut microbiota species

Among the strongest findings with species-level annotation, MPA was associated with higher abundance of *P. copri* and *F. prausnitzii* (HG3A.0241 and HG3A.0025). Time spent in exercise has previously been associated with higher *P. copri* abundance in cyclists.^37^ Nevertheless, the associations between *P. copri* and health outcomes have been inconsistent, with some studies suggesting a role in fibre fermentation and improved glucose metabolism,^38^ while others reported associations with insulin resistance^39^ and hypertension.^40^ MPA and VPA had similar associations, but there were some clear exceptions. MPA was associated with higher abundance of *P. copri* and *F. prausnitzii* (HG3A.0025), and lower abundance of *B. uniformis* and *P. vulgatus*, while we could not detect the same associations for VPA. We found that VPA was positively associated with *Roseburia homininis* and *F. prausnitzii* (HG3A.0241), but negatively associated with another subspecies of *F. prausnitzii* (HG3A.0010). In general, *F. prausnitzii* is considered to have anti-inflammatory properties^41^ and individuals with type 2 diabetes have been reported to have a lower abundance of this bacteria.^42^ A higher abundance of *F. prausnitzii* and *R. hominis* has also been reported in active premenopausal women, based on accelerometer assessment.^43^ Both *F. prausnitzii* and *R. hominis* are important producers of butyrate, which is a main energy source for colonocytes and critical for gut homeostasis.^44^ A previous study in a small sample of young adults has also reported an association between cardiorespiratory fitness and fecal butyric acid.^45^

In the present study, SED was associated with higher abundance of *B. obeum* (HG3A.0001 and HG3A.0009), *R. torques*, *P. vulgatus,* and *Escherichia coli*. An increased abundance of the *Escherichia/Shigella* taxon was also reported in sedentary individuals in the Healthy Life in an Urban Setting cohort (HELIUS, N = 1334), the largest population-based study on physical activity and gut microbiota until the present study.^22^ A higher abundance of *E. coli* has been observed in individuals with atherosclerotic cardiovascular disease^46^ and users of the anti-diabetes medication metformin,^47^ suggesting a link between physical activity, gut microbiota, and cardiometabolic diseases. Due to different microbiota profiling methods, our findings cannot be directly compared with the HELIUS study. We confirmed though the positive association between *P. copri* and physical activity but found a positive association between VPA and *R. hominis*, which was more abundant in sedentary individuals in the HELIUS study.

The median VPA in our study sample was only 0.06%. A previous study has suggested that as little as 15 min/week of VPA reduces the risk for all-cause mortality by 18%;^48^ therefore, effects of VPA could be expected even with little time spent in these activities. Vigorous-intensity activities include running and high-intensity sports, while moderate-intensity activities cover a wider range of activities, including brisk walking and bicycling at low speed, but also household chores.^49^ Future studies could further investigate the associations of subcategories of MPA with the gut microbiota.

### Adjustment for BMI and WHR had a pronounced effect on the associations observed between SED and species of the gut microbiota

In the models additionally adjusted for dietary variables or for BMI/WHR, we observed attenuated coefficients. The largest effect was observed for associations with SED after adjustment for BMI/WHR. This attenuation could be caused by BMI acting as a confounder or a mediator. However, for 14 species, including *F. prausnitzii* (HG3A.0025), the association with SED became stronger after BMI/WHR adjustment. This could be due to a negative confounding effect of adiposity, which masked the true associations in the main model, or that the adjustment for BMI/WHR led to collider bias. The greater impact of adjusting for BMI/WHR on SED coefficients than on MPA or VPA coefficients aligns with previous Mendelian randomization analyses, which have demonstrated stronger bidirectional effects between SED and BMI than between physical activity time and BMI.^50,51^

### Sedentary behaviour associated with higher gut microbiota capacity to degrade amino acids and to produce certain short-chain fatty acids

With regards to the functional potential of the gut microbiota, we found that SED was associated with lower capacity for fibre degradation and higher capacity for amino acid degradation. Likewise, a previous study has reported an increased abundance of carbohydrate degradation pathways in athletes.^52^ These findings could be due to lower fibre content in the diet of sedentary individuals, which we aimed to address by adjusting for fibre intake. A lower availability of fermentable carbohydrates in the distal gut can result in a reduction in saccharolytic bacteria and an increase in proteolytic bacteria.^30^ Intervention studies with standardized diet would be needed to disentangle physical activity associations from associations due to differences dietary intakes.

In our main model results, MPA was negatively associated with modules for short-chain fatty acids synthesis, more specifically acetate and propionate synthesis, while SED was positively associated with these modules. Conversely, six weeks of exercise training has been reported to increase the abundance of bacterial genes involved in propionate synthesis.^53^ The same study also reported a BMI-dependent exercise-induced increase in bacterial genes of butyrate synthesis. In our study, VPA was positively associated with butyrate synthesis in the main model, while in the BMI/WHR-adjusted model, it was SED that was positively associated with this same module. For other modules in the microbiota-gut-brain axis, SED was negatively and MPA positively associated with the module for the heat-shock protein ClpB that has been suggested to influence appetite regulation.^54^ SED was also associated with higher abundance of the GABA synthesis module and higher abundance of *E. coli*, one of the main GABA-producing bacteria in the gut.^55^ Although lower plasma levels of GABA have been described in individuals with depression,^56^ recent studies found elevated levels in a mixed sample of medicated and non-medicated individuals with major depressive disorder.^57,58^

### Strengths and limitations

The strengths of this study are the accelerometer-based assessment of sedentary behaviour and physical activity phenotypes, the large sample of participants from the general population, and the high taxonomic resolution microbiome data. Moreover, we had access to comprehensive information on potential confounders. Some limitations apply. One concern is whether the associations described reflect the lifestyle of health-conscious individuals. Despite covariate adjustments, it is implausible to capture all dimensions of dietary intake and residual confounding may remain. Accelerometers measure absolute physical activity intensity, but relative intensity could be more clinically relevant. Standardized accelerometer cut-offs can misclassify low and high fitness individuals. Estimating relative intensity would though require data on individual maximal capacity, such as maximal oxygen uptake.^59^ Additionally, social desirability bias and adherence to the study instructions could affect accelerometer-based assessment. However, these misclassifications would be non-differential. The accelerometer needed to be removed during water-based activity and may also underestimate physical activity intensity during cycling, upper body activities, and weight-lifting.^25^ Our study was conducted in a Swedish population aged 50-65, thus generalizability to other populations is limited. Additionally, we cannot assess the direction of the associations using cross-sectional data. It is suggested that gut microbiota may favour the practice of exercise by enhancing the enjoyment of physical activity^60^ or improving the host performance.^61^ The gut microbiota can also affect adiposity,^62^ which is negatively associated with physical activity. Lastly, the use of relative abundances incur on the issues of compositional data, which can produce false-positive associations.^63^

In summary, sedentary behaviour and physical activity were associated with a large number of gut microbiota species and functional modules. Our findings can be used to guide research on the interplay between physical activity, the gut microbiota composition, and health outcomes.

## Data Availability

The de-identified metagenomic sequences can be found in the European Nucleotide Archive under the accession code PRJEB51353. The statistical analyses R codes can be found at https://github.com/MolEpicUU/physicalactivity_gut. The individual-level data underlying this article were provided by the SCAPIS study and are not shared publicly due to confidentiality. Data will be shared upon reasonable request to the corresponding author only after permission by the Swedish Ethical Review Authority (https://etikprovningsmyndigheten.se) and by the SCAPIS Data access board (https://www.scapis.org/data-access/).

## Contributorship

JGS, GBergström, LL, GE, JÄ, MO-M and TF obtained the funding for the study. GBaldanzi, SS-B, EEB, ÖK, UH, BK, and TF planned and designed the study. GBaldanzi carried out the statistical analyses with contribution from SS-B, KFD, and UH. GBaldanzi wrote the first version of the manuscript with support from SS-B, BK, and TF. All authors contributed with the critical interpretation of the results and revision of the manuscript.

## Funding/support

Financial support was obtained in the form of grants from the European Research Council [ERC-STG- 2018-801965 (TF); ERC-CoG-2014-649021 (MO-M); ERC-STG-2015-679242 (JGS)], the Swedish Heart-Lung Foundation [Hjärt-Lungfonden, 2019-0505 (TF); 2018-0343 (JÄ); 2020-0711 (MO-M); 2020-0173 (GE); 2019-0526 (JGS)], the Swedish Research Council [VR, 2019-01471 (TF), 2018-02784 (MO-M), 2019-01015 (JÄ), 2020-00243 (JÄ), 2019-01236 (GE), 2021-02273 (JGS), 2018-02837 (EXODIAB 2009-1039)], the Swedish Research Council for Sustainable Development [FORMAS, 2020- 00989 (SA)], EASD/Novo Nordisk (SA), Göran Gustafsson foundation [2016 (TF)], Axel and Signe Lagerman’s foundation (TF), the A.L.F. governmental grant 2018-0148 (MO-M), The Novo Nordic Foundation NNF20OC0063886 (MO-M), The Swedish Diabetes Foundation DIA 2018-375 (MO-M), the Skandia Risk&Hälsa, 2023 (ÖE, EEB), and governmental funding of clinical research within the Swedish National Health Service (JGS). The study was also supported by the Swedish Foundation for Strategic Research (LUDC-IRC 15-0067). Funding for the SCAPIS study was provided by the Swedish Heart-Lung Foundation, the Knut and Alice Wallenberg Foundation, the Swedish Research Council and VINNOVA (Sweden’s Innovation agency), the University of Gothenburg and Sahlgrenska University Hospital, Karolinska Institutet and Stockholm County council, Linköping University and University Hospital, Lund University and Skåne University Hospital, Umeå University and University Hospital, Uppsala University and University Hospital.

## Competing interests

J.Ä. has served on the advisory boards for Astella, AstraZeneca and Boehringer Ingelheim and has received lecturing fees from AstraZeneca and Novartis, all unrelated to the present work. Remaining authors declare no competing interests.

## Acknowledgments

We acknowledge the SCAPIS board for enabling the current study, along with the Swedish Heart-Lung Foundation, the main funding body of SCAPIS. The computations and data handling were enabled by resources in project sens2019512 provided by the National Academic Infrastructure for Supercomputing in Sweden (NAISS) and the Swedish National Infrastructure for Computing (SNIC) at Uppsala Multidisciplinary Center for Advanced Computational Science (UPPMAX), funded by the Swedish Research Council through grant agreement no. 2018-0597 and no. 2022-06725.

## Data Sharing

The de-identified metagenomic sequences can be found in the European Nucleotide Archive under the accession code “PRJEB51353”. The statistical analyses R codes can be found at https://github.com/MolEpicUU/physicalactivity_gut. The individual-level data underlying this article were provided by the SCAPIS study and are not shared publicly due to confidentiality. Data will be shared upon reasonable request to the corresponding author only after permission by the Swedish Ethical Review Authority (https://etikprovningsmyndigheten.se) and by the SCAPIS Data access board (https://www.scapis.org/data-access/).

## Notes

### Author Declarations

The Swedish Ethical Review Authority approved the Swedish CardioPulmonary bioImage Study (DNR 2010-228-31M) and the present study (DNR 2018-315 with amendment 2020-06597). All participants provided written informed consent.

