## Supplemental figures 1-3 for "ACCELEROMETER-BASED SEDENTARY BEHAVIOUR AND PHYSICAL ACTIVITY ARE ASSOCIATED WITH THE GUT MICROBIOTA IN 8507 INDIVIDUALS FROM THE POPULATION-BASED SCAPIS"

Supplemental Figure 1

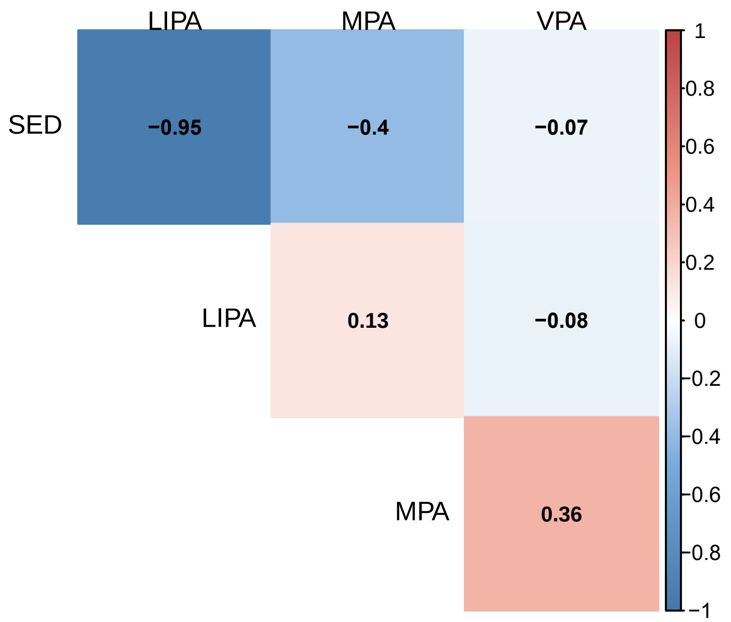


Supplemental Figure 1. Spearman’s correlation between the accelerometer-based variables. SED: percentage of time in sedentary behavior; LIPA: percentage of time in low-intensity physical activity; MPA: percentage of time in moderate-intensity physical activity; VPA: percentage of time in vigorous-intensity physical activity.

Supplemental Figure 2

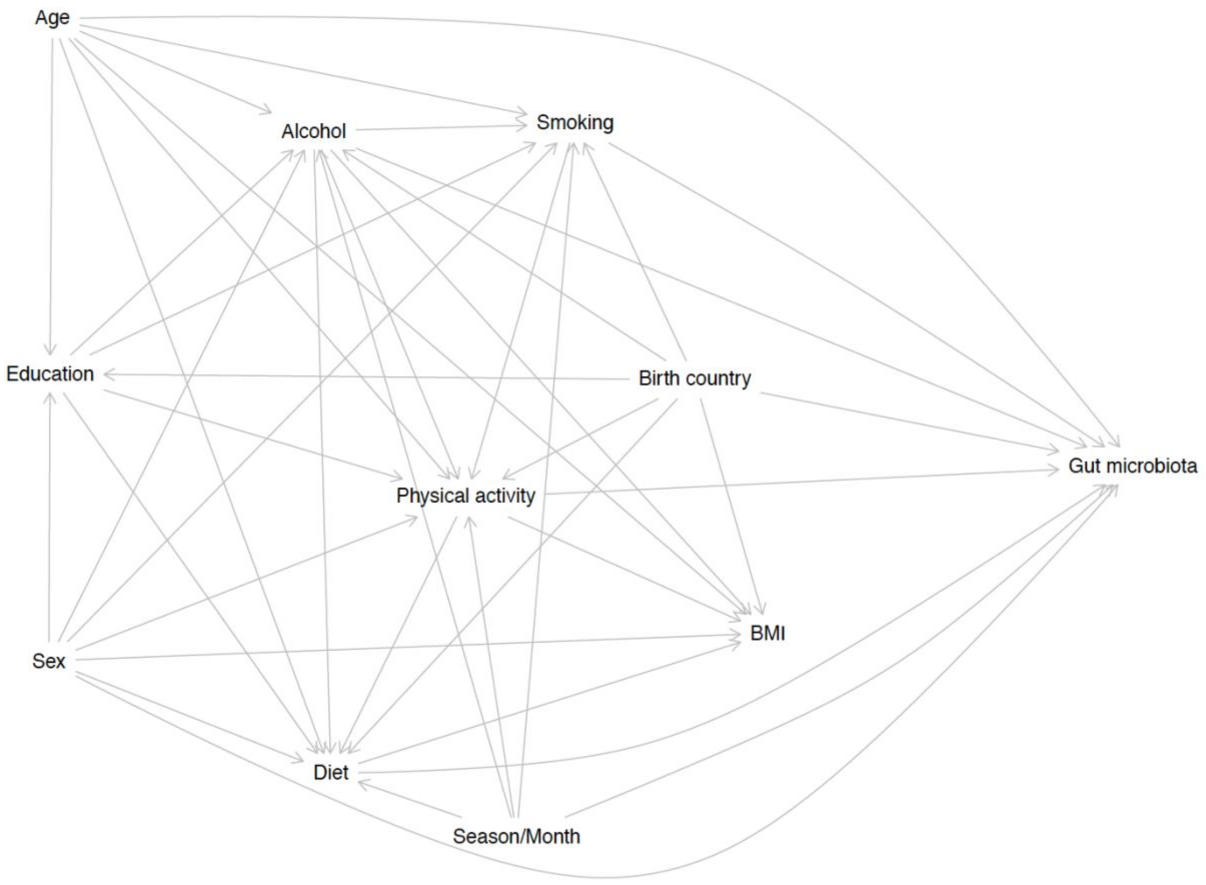


Supplemental Figure 2. Directed acyclic graph depicting the assumptions about the association between physical activity and the gut microbiota.

Supplemental Figure 3

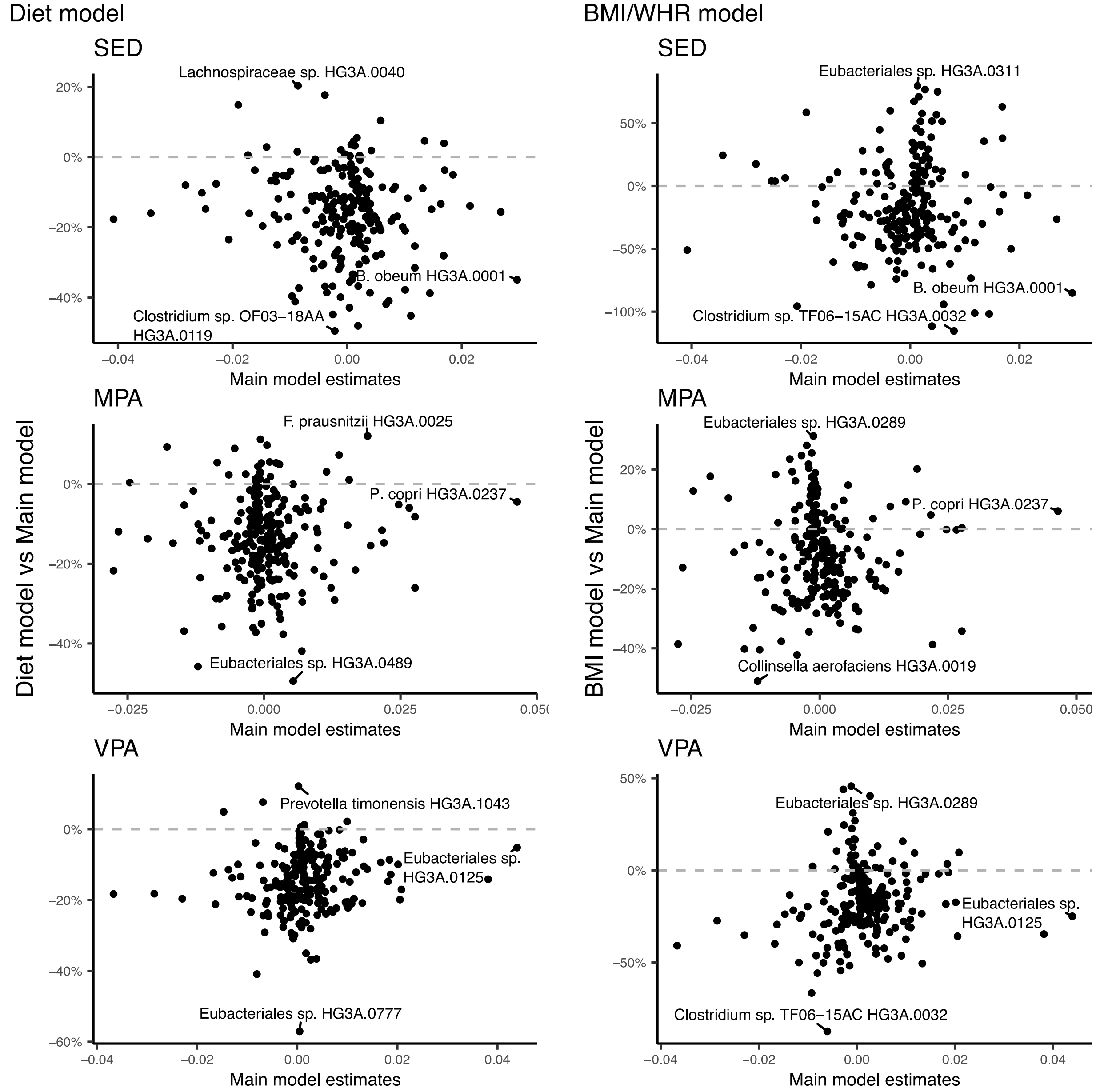


Supplemental Figure 3. Changes in the regression coefficients from the main model after adjustment for the diet model or for the BMI/WHR model. The x-axis shows main model estimates and the y-axis shows the percentual change to the main model coefficients after the additional adjustments. Here presented are the species associated in the main model after multiple testing adjustment (False discovery rate of 5%). Main model: adjustment for age, sex, alcohol intake, smoking, education, country of birth, study site, month of accelerometer wear, total accelerometer wear time, percentage of wear time on weekend, and fecal DNA extraction plate. Diet model: main model with additional adjustment for total energy intake, and percentage of energy intake from carbohydrates, protein, fibers, and added sugars. BMI/WHR model: main model with additional adjustment for BMI and waist-hip ratio. SED: percentage of time in sedentary behavior; MPA:

percentage of time in moderate-intensity physical activity; VPA: percentage of time in vigorous-intensity physical activity.
