## Supplemental methods for "ACCELEROMETER-BASED SEDENTARY BEHAVIOUR AND PHYSICAL ACTIVITY ARE ASSOCIATED WITH THE GUT MICROBIOTA IN 8507 INDIVIDUALS FROM THE POPULATION-BASED SCAPIS"

#### Accelerometer data processing

Detailed information on the accelerometry data processing in SCAPIS has been published by Ekblom-Bak et al.[1] In this study, we applied the same protocol with the refinement of excluding registrations during estimated bedtime. Participants were instructed to wear the ActiGraph tri-axial accelerometer (GT3X+, wGT3X+ or wGT3X-BT, Actigraph LCC, Pensacola, USA) over the hip for seven consecutive days during the whole day, except during sleep and water-based activities. Despite the instructions to remove the hip-worn accelerometer during sleep, 1480 participants (1319 from Uppsala) had unreasonably high average daily accelerometer wear time; i.e., >18h/day. This indicates that many participants did not remove the accelerometer during sleep. Therefore, we used information from a second accelerometer, that was attached to the thigh, concomitantly to the hip-worn accelerometer, used by SCAPIS-Uppsala participants only. The thigh accelerometer data from 3780 Uppsala participants was processed with the custom-made software ActiPASS 1.42 (www.github.com/Ergo-Tools/ActiPASS). ActiPASS uses the rotation and angle of the thigh in relation to the gravity line, to identify the time in prolonged lying bouts.[2] This was used as a proxy for bedtime. Based on this information, we filtered the hip-worn accelerometer data of all Malmö and Uppsala participants to registrations between 05:52 a.m. and 11:46 p.m. in weekdays and 07:15 a.m. and 11:59 p.m. in weekends, which corresponded to the first quartile of bedtime end and the third quartile of bedtime start based on thigh-accelerometer data.

The raw hip-worn accelerometry data was transformed into counts per minute (cpm) over 60s epochs. Sedentary time was defined as <200 cpm, low-intensity physical activity as 200 – 2689 cpm, moderate-intensity physical activity as 2690 – 6166 cpm and vigorous as ≥6167 cpm.[1] Periods of ≥60 min with cpm equal to zero, with intervals of maximum two minutes of 0 – 199 cpm, were defined as non-wear time. A valid day was defined as a day with >10 hours of wear time and we excluded 467 participants with <4 valid days. The percentage of time in sedentary (SED), in low-intensity (LIPA), in moderate-intensity (MPA), or in vigorous-intensity (VPA) physical activity were calculated as the time spent in these activities divided by the total wear time.[1]

#### Fecal metagenomics

The gut microbiota analysis processing has been previously described.[3] In summary, fecal samples were collected at home using the material provided at the first visit to the test center. Participants were instructed to keep samples at the home freezer until the second visit to the study site, for further shipping to the central biobank for storage at -80°C. From the biobank, the sample boxes were shipped on ice in random order to Clinical Microbiomics A/S (Copenhagen, Denmark) for processing during 2019 and 2020.

DNA extraction was performed using NucleoSpin® 96 Soil kits (740787; Macherey-Nagel; Germany, batch number. 1903/001). Library preparation used the NEBNext® DNA Library Prep Kit (New England Biolabs). Sequencing was performed with Ilumina Novaseq 6000 (Illumina, USA) and generated an average read count of 25.3 million (SD = 3.8 million) read-pairs for Uppsala samples and 26.8 million (SD = 6.9 million) read-pairs for Malmö samples. Reads with adapter contamination, >10% ambiguous bases, >50% bases with Phred score <5, and reads mapped to human reference genome GRCh38 were removed. Reads were them mapped to metagenomic species as defined by Clinical Microbiomics A/S[4] and species counts were normalized according to the read length. Taxonomic annotation was based on the NCBI RefSeq database (downloaded on 2 May 2021). A downsized or rarefied species count matrix was also produced by random sampling, without replacement, from the subset of gene count table corresponding to the metagenomic species signature genes. The final read count for the downsized data was 210 430 read-pairs. One sample had significantly less reads assigned to the metagenomic species signature genes and was not included in the downsized matrix. The downsized matrix was used to calculate the alpha and beta-diversity indices. All other analyses were performed with the non-downsized data.

The functional potential of the gut microbiota was defined in terms of gut metabolic modules[5] (GMM) and microbiota-gut-brain modules[6] (MGB). To calculate the modules abundance, reads were mapped to Kyoto Encyclopedia of Genes and Genomes (KEGG) orthology (KO) database (https://www.genome.jp/kegg/) using EggNOG-mapper-software (v. 2.0.1).[7] We removed one of the modules when there was an overlapped between GMM and MGB, thus we removed the modules MGB049, MGB 036, MF0030, MF0031, MF0032, MF0047, MF0075, MF0076, MF0082, MF0088, MF0089, MF0094, MF0095. Next, we used Omixer-RPM v.0.3.2[8] considering a minimum module coverage of 66.6%. We focused on species, GMMs, MGBs with a relative abundance > 0.01% in ≥1% of the individuals, resulting in 1336 species, 76 GMMs, and 28 MGBs for further analyses. For 12 samples, it was not possible to perform the fecal metagenomics (e.g. low DNA yield) and one sample had a low number of mapped reads. These 13 participants were excluded.

### Covariates

Covariate information was obtained from the SCAPIS questionnaire, and anthropometric measurements and fasting plasma samples collected during a study site visit. From the food frequency questionnaire, the mean daily intake of alcohol, fiber, added sugar, protein, carbohydrate, fat (all g/day), and total energy (kcal/day) were estimated.[9,10] Added sugar intake was estimated by summing the intakes of sucrose and monosaccharides, followed by subtraction of the calculated mean intake of those sugars from common Swedish fruits and vegetables including juices (sucrose intake + fructose intake + glucose intake - fruit and berry intake×0.10 - vegetable intake×0.03 -juice intake×0.08).[11] Protein, carbohydrate, fat, fiber, and added sugar intake were transformed to percentages of non-alcohol energy intake. Women with an estimated average energy intake ≤500 or ≥5000 kcal/day and men with an energy intake ≤550 or ≥6000 kcal/day were considered over or under-reporters of dietary intake and their dietary variables were set to missing. We categorized smoking status as current, former, or non-smoker, and highest achieved education level as incomplete compulsory education, complete compulsory education, secondary education, or university education. Country of birth was grouped as Scandinavia (Sweden, Denmark, Norway, or Finland), non-Scandinavian Europe, Asia, and other countries. For the variable "month when the accelerometer was worn" the categories June and July were merged because there were only 2 participants for July.

Data on dispensed prescribed medications were obtained from the Swedish Prescribed Drug Register held by the National Board of Health and Welfare, which includes information on all pharmacological agents prescribed in Sweden since 2005.[12] We retrieved from the drug register information on medications prescribed within six months before the first study site visit for hypertension (Anatomical Therapeutic Chemical codes [ATC] C02, C03A, C03EA01, C08C, C09, and C07 – except C07AA07 sotalol),[13] diabetes (ATC A10),[14] dyslipidemia (ATC C10), depression (ATC N06A), and anxiety (ATC N05B). If a participant reported in the questionnaire to have used a medication for hypertension, diabetes, or dyslipidemia in the last two weeks, the participant was defined as a user for that medication even if a prescription had not been dispensed in the last six months. Because proton pump-inhibitors are allowed to be bought without a prescription in Sweden, proton-pump inhibitor usage was defined as a measurable level of omeprazole or pantoprazole metabolites in the plasma.[3] Antibiotic use was not present in the questionnaire data. Thus, antibiotic use was defined as a dispensed prescription (ATC code J01) up to three months before the first study site visit.
